# Adapting COVID-19 research infrastructure to capture Influenza and Respiratory Syncytial Virus alongside SARS-CoV-2 in UK healthcare workers Winter 2022/23 and beyond: protocol for a pragmatic sub-study

**DOI:** 10.1101/2023.09.19.23295789

**Authors:** Jonathan Broad, Dominic Sparkes, Naomi Platt, Anna Howells, Sarah Foulkes, Jameel Khawam, Michelle Cole, Nick Andrews, Conall Watson, Susan Hopkins, Victoria Hall, the SIREN study team

**Author notes:** Joint first authors (Jonathan Broad, Dominic Sparkes, Naomi Platt).

## Abstract

**Introduction:** During the COVID-19 pandemic, extensive research was conducted on SARS-CoV-2, however important questions about other respiratory pathogens remain unanswered. A severe influenza season in 2022-2023 with simultaneous circulation of SARS-CoV2 and Respiratory Syncytial Virus (RSV) is anticipated. This sub-study aims to determine the incidence and impact of these respiratory viruses on healthcare workers (HCW), the symptoms they experienced, the effectiveness of both COVID-19 and influenza vaccination and the burden of these infections on the National Health Service (NHS) workforce.

**Methods and analysis:** This is a longitudinal prospective cohort sub-study, utilising the population and infrastructure of SIREN, which focuses on hospital staff in the UK. Participants undergo fortnightly Nucleic Acid Amplification Testing (NAAT) on a multiplex assay including SARS-CoV-2, Influenza A&B and RSV, regardless of symptoms. Questionnaires are completed every two weeks, capturing symptoms, sick days, exposures, and vaccination records. Serum samples are collected monthly or quarterly from participants associated with a SIREN site. This sub-study commenced on 28/11/22 to align with the predicted influenza season and participants’ influenza vaccine status. The SIREN Participant Involvement Panel (PIP) shaped the aims and methods for the study, highlighting its acceptability. UK Devolved Administrations were supported to develop local protocols. Analysis plans include incidence of asymptomatic and symptomatic infection, comparisons of vaccination coverage; assessment of sick day burden, and effectiveness of seasonal influenza against infection and time off work. Data are also integrated into UKHSA nosocomial modelling.

**Ethics and dissemination:** The protocol was approved by the Berkshire Research Ethics Committee (IRAS ID 284460, REC Reference 20SC0230) on 14/11/2022. Participants were informed in advance. As the frequency and method of sampling remained the same, implied consent processes were approved by the committee. Participants returning to the study give informed consent.

Regular reports to advisory groups and peer-reviewed publications are planned to disseminate findings and inform decision making. **Trial registration number: ISRCTN11041050.**

**Strengths and limitations of this study:**

- The positioning of this sub-study, led by public health agencies in collaboration with a network of NHS sites, facilitates horizon scanning enabling rapid adaptation of the study protocol and deployment to conduct relevant scientific research in a cohort of healthcare workers
- To achieve target recruitment, deployment of a new postal pathway is underway, allowing for more direct communication between the central research team and participants
- The multi-disciplinary partnerships including a network of academic centres established and embedded by SIREN can now be leveraged and extended to explore ‘Flu and RSV
- Decentralised study delivery, with testing at a network of NHS sites has both strengths and limitations. This includes enabling more in-depth relationships and communications between participants and their local research teams, however, introduce additional communication, governance, and data sharing requirements.
- Some demographics are over-represented such as female staff, nurses and doctors, and some are under-represented such as staff from ethnic minorities, porters and estates. This is partly the consequence of a rapid recruitment drive at the beginning of the COVID-19 pandemic.

## Background and rationale

The COVID-19 pandemic has caused significant global morbidity and mortality.^1^ Despite the success of vaccinations and medical interventions in reducing severe disease, COVID-19 is now endemic, leading to a shift in governmental policy towards “living with COVID-19”.^2^ As a result, SARS-CoV-2 co-exists with other respiratory pathogens, including Influenza and Respiratory Syncytial virus (RSV), both of similar public health importance with epidemic potential. These viruses contribute to waves of illness, hospitalisation and death across all ages but particularly, in the case of RSV, infants and the elderly.^3-5^ Relaxation of non-medical interventions such as social distancing, in Australasia during Winter 2022 resulted in a significant rise in Influenza cases, hospital admissions and deaths, compared to previous seasons.^6^ Surveillance data from the UK showed an early start to the 2022/23 Influenza season, alongside persistently high rates of RSV throughout the pre-winter months.^7 8^ Understanding the incidence and impact of these respiratory viruses is required for healthcare and workforce planning.

Many research questions about the manifestation, natural history and vaccine effectiveness of influenza and RSV remain answered, despite progress with SARS-CoV-2. Incidence rates of both infections is skewed towards symptomatic illness detection, given nationwide screening has not been feasible in the same way as for SARS-CoV-2, for example in healthcare workers, travellers and hospital admissions. Published rates of asymptomatic influenza derive mostly from outbreak investigations and household transmission studies, which may not differentiate between truly asymptomatic and pre-symptomatic cases.

Healthcare workers are an important population to study given their high exposure to respiratory pathogens and the impact of infection on workforce resilience and nosocomial transmission dynamics.^9 10^ Their access to testing facilities and their commitment to medical and scientific research make them a feasible population to study.

The SARS-CoV-2 Immunity & Reinfection Evaluation (SIREN) study is a prospective cohort study of healthcare workers (HCWs) across the UK.^11^ It was initiated early in the pandemic to evaluate the risk of reinfection with SARS-CoV-2 and has adapted its objectives to the evolving pandemic.^12^ Given the threat of a triple-pandemic of SARS-CoV-2, Influenza and RSV in winter 2022/23, the SIREN study team rapidly initiated a protocol for the described sub-study to introduce multiplex PCR testing in the cohort, testing for Influenza and RSV alongside SARS-CoV-2. This provides a valuable opportunity to investigate under-researched aspects of influenza and RSV, including incidence of asymptomatic influenza and RSV in healthcare workers, the sick day burden and influenza vaccine effectiveness in a healthy working aged population.

## Methods and analysis

### Study design

This is a prospective longitudinal cohort sub-study, within the larger SIREN study. Sites and participants are recruited into the Winter Pressures sub-study from October 2022 until February 2023. Individuals are followed up until 31 March 2023. The planned study set up of timelines is based around a notional Influenza season start date of 28^th^ November, based on previous seasonal peaks, although the actual start date is based on pragmatic limitations of adapting an ongoing cohort study.

### Study objectives

The overall aim of this sub study is to investigate the burden of respiratory viruses, alongside SARS-CoV-2, experienced by UK healthcare workers over the winter season 2022/23.

Primary objectives:

- To estimate the incidence of influenza and RSV infection in HCWs using regular PCR testing.
- To estimate the effectiveness of 2022/23 seasonal Influenza vaccination against Influenza infection in HCWs.

Secondary objectives:

- To estimate the incidence of symptomatic and asymptomatic COVID-19, Influenza and RSV infection.
- To compare symptom reporting to identify potential symptom variation between respiratory pathogens
- To estimate the incidence of co-infection with SARS-CoV-2, influenza and RSV.
- To estimate the number of days off work associated with respiratory illness over winter 2022/23.
- To monitor the serological response to influenza vaccination over time.

## Participants and recruitment

### Population and eligibility

Winter Pressure sub-study participants are recruited from the SIREN cohort of staff members at NHS healthcare organisations across the four nations of the UK. Anyone that is eligible for SIREN is eligible for the sub-study. The eligibility criteria for the 44,000 SIREN participants recruited between June 2020 and March 2021 was: aged over 18-years, working at an NHS organisation participating in SIREN, able to participate in active testing and willing to remain engaged in follow-up.

### Study visits

Participants are invited to take fortnightly swab tests, either by post or at the clinic according to their pathway (figure 1). Participants are invited to complete fortnightly online surveys on their electronic devices. Participants are invited to take monthly or quarterly blood tests according to capacity at their research site.

**Figure 1:**
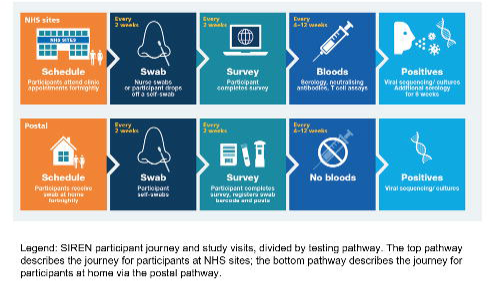
Diagram outlining the SIREN participant journey.

### Recruitment and consent

Participants are recruited to the winter pressures sub-study by two routes: via their SIREN site if their site joins the Winter Pressures pathway or directly by the SIREN study team to join a new Winter Pressures postal testing sub-study (figure 2).

**Figure 2:**
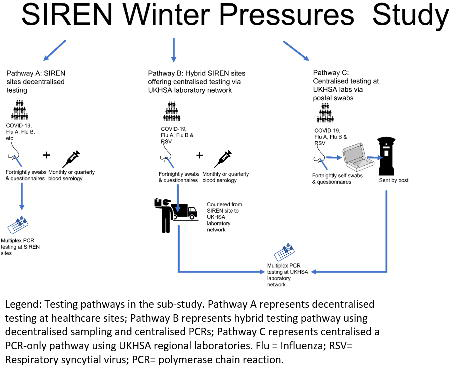
Description of the SIREN Winter Pressures sub-study testing pathways.

#### Recruitment of sites

#### Recruitment of participants at sites

From October 2022 onwards, SIREN sites are actively testing participants (a total of 66 sites), are invited to join the sub-study. To participate, sites must have the capacity to switch PCR testing from a monoplex PCR platform to a multiplex PCR platform, covering at least SARS-CoV-2 and Influenza. For a few sites without local access to multiplex PCR testing, an alternative testing pathway is offered (figure 2).

This site-based model ensures that all participants under active follow up at Winter Pressures sites are tested on multiplex PCR platforms throughout winter 2022/23.

Regular dialogue through webinars and newsletters has kept participants informed about the planned study, and sites have been encouraged to engage with participants about their involvement. Active participants at winter pressures sites are informed in advance that their samples will now be tested using multiplex PCR tests for COVID-19, Influenza and possibly RSV, with communication of any local implications handled by respective sites. The UKHSA SIREN team send direct individual communications, including the participant information leaflet (Appendix 4-6). Participants who do not wish to have their samples tested on the multiplex PCR platform are able to ‘opt out’ using the study withdrawal process. Explicit re-consent is not sought and by providing a sample after the receipt of the PILS they consent to joining the study based on guidance from the HRA Ethics Committee.

To increase participant numbers, participants at Winter Pressure sites who are not actively being followed-up, are invited again. They are informed that their samples will be tested on multiplex platforms. Participants receive one invitation message via email and /or SMS according to their preference, followed by two reminder messages.

#### Recruitment of participants from closed out sites to new postal testing pathway

In recognition of the inability of some SIREN sites to offer multiplex, a centralised postal testing pathway has been developed (figure 1). Eligible participants for this pathway are those who have not withdrawn from the study but are no longer in active testing, due to the closure of their SIREN site. The UKHSA SIREN team directly recruits participants through electronic invitations including a Winter Pressures sub-study participant information leaflet sent via email and/or SMS (Appendix 4-6). Participants receive a link to an electronic consent form.

Recruitment to the postal pathway takes place between 12/12/2022 and 23/01/2023. Whilst reflecting the pragmatic challenges of setting up a centralised testing process. To improve cohort diversity, eligible participants from underrepresented groups in the main cohort are invited first and have two days to enrol before invitations are sent to the remaining eligible participants. Underrepresented groups include porters and estates staff, males and participants from ethnic minority backgrounds, as reported in their original SIREN baseline questionnaire. Recruitment will be closed by 23/01/2023 or once 5000 participants have been recruited, whichever is sooner.

#### The NIHR Clinical Research Network

The SIREN study is an NIHR prioritised study and when designing the Winter Pressures sub-study regular communication was made with the NIHR Clinical Research Network (CRN) who offered support. As a mechanism of promoting engagement from sites accruals were offered for each participant who was recruited to the winter pressures sub-study. Sites could therefore claim an accrual for any participant who consented to join.

## Data collection and data management

### Sampling and laboratory testing

There are three testing and sampling pathways in the SIREN Winter Pressures sub-study (figure 2):

1. Site-based testing using multiplex COVID-19,Influenza and potentially RSV PCR and serology in addition to current protocol;
2. Postal pathway testing using multiplex PCR only via centralised laboratory;
3. Hybrid NHS site-based testing and centralised referral of samples to UK Health Security Agency (UKHSA) regional laboratories.

#### Site based PCR testing

Participants in the core SIREN sites pathway provide nose and throat swabs via self-swabbing or clinician-swabbing on a fortnightly basis, in addition to monthly or quarterly venous sampling of up to 10mls of blood by a trained phlebotomist. Multiplex PCR platforms vary by site, but sites were required to have at least SARS-CoV-2, Influenza A, and Influenza B viruses included.

#### Postal pathway PCR testing

Participants in the postal pathway are provided with packs received via the post which contain a self-swab kit, comprised of a nose swab and throat swab, a 3ml vial of VTM for the 2 swabs to be placed into, a container for the vial, 2 plastic bags and UN3373 compliant packaging with a Track24 Return label attached. An instruction leaflet was provided to explain the process (Appendix 7). To fulfil data linkage requirements for uploading to the Secondary Generation Surveillance System (SGSS) two matching unique barcodes were provided, with one to be stuck to the vial and the other to a request form which also included space for name, StudyID and date of birth as well as collection date, to ensure there were 3 unique identifiers for linkage (see data linkage below). See Appendix 2 for the complete UKHSA regional laboratory process. The UKHSA regional laboratories utilise the Hologic Panther Fusion Multiplex assay, which tests for SARS-CoV-2, Influenza A/B, and RSV. For swabs tested at the UKHSA regional laboratories, in addition to arrangements for sequencing SARS-CoV-2 positives as per the main study protocol, VTM was retained from samples positive for Influenza and RSV and biobanked centrally for potential future sequencing. Participants in the postal pathway did not provide any serology.

#### Hybrid NHS site-based testing and centralised referral of samples

Participants in the hybrid pathway provided monthly or quarterly venous sampling of up to 10mls of blood by a trained phlebotomist. They also provided nose and throat swabs to their SIREN site.

These samples were tested locally for SARS-CoV-2 but referred to the UKHSA regional laboratories for multiplex testing. There were two pathways used by sites: 1) participants provided 2 vials with swabs – with 1 vial sent to the local laboratory and 1 sent to the regional laboratory. 2) participants provided 1 vial to their local laboratory which was tested for SARS-CoV-2 and the residual VTM was sent to the UKHSA regional laboratories for multiplex testing. Samples were referred using a dedicated request form and line list and sent via courier to the UKHSA laboratories (see Appendix 3 for the flow chart and referral forms). The testing methodology at the UKHSA laboratories was the same as outlined above.

#### Serology

Longitudinal serum samples collected at sites from SIREN participants are biobanked centrally within the UKHSA Seroepidemiology Unit (SEU), and new samples collected from participants enrolled in the Winter Pressures sub-study at sites are added to this repository. These samples can be accessed and tested to investigate antibody responses to Influenza (including vaccination) and RSV using assays available within UKHSA and via academic partners.

### Follow up questionnaire

Every participant regardless of their pathway completed the same questionnaire, an adaptation of the SIREN study follow-up questionnaire. Additional questions have been added to capture influenza vaccination, including the date and type of vaccine. The number of “sick days”, i.e., time taken off work due to a respiratory illness were also captured, along with exposures to respiratory viruses at work and whether appropriate (as defined by their workplace) personal protective equipment was worn. See appendix 1 for the full questionnaire

### Data management

Laboratories in England are required to notify COVID-19 and Influenza results (positives and negatives) via the Second-Generation Surveillance System (SGSS) on a daily basis. Similar systems are established in the public health bodies of the devolved administrations, including SAIL database and the Electronic communication of Surveillance in Scotland. For RSV, laboratories only report positive results, so these are combined with SARS-CoV-2 and influenza, where negative results are reported, to obtain a denominator.

The SIREN study team utilises deterministic linkage by NHS number, StudyID, name or centrally assigned specimen number (available on barcodes) to obtain results from SGSS. Results from the devolved administrations are pulled from their respective databases in a similar manner by their public health bodies and securely transferred to the central SIREN study team for inclusion in the SIREN database.

Participants in the postal pathway are asked to complete their fortnightly questionnaire on the same date as the swab provide their barcode number for additional linkage.

Vaccination coverage data for both COVID-19 and Influenza vaccine date and type was collected via the National Immunisation Management System (NIMS) in England, and equivalent databases in the devolved administration, and also collected as part of the follow up questionnaire.

### Sample size and analysis plan

Target recruitment for the Winter Pressures sub-study is determined by a sample size calculation for estimating the effectiveness of the 2022/23 seasonal vaccination against Influenza infection. The assumptions were: Influenza PCR positivity per week of 0.5 % (low scenario), 1% (medium), 1.5% (high); influenza PCR positivity lasts 1 week, a 14-week influenza season, and vaccination coverage at 60% based on the previous 2 years in this cohort. Using a test-negative case control (TNCC) method, a sample size of 7500 participants would offer sufficient precision with estimated confidence intervals around an observed VE of 50% of 34-62 if weekly positivity is 0.5%, 39-62 if weekly positivity is 1.0%, and 41-57 if weekly positivity is 1.5%.

The Vaccine effectiveness analysis will be based on TNCC and also a survival analysis to estimate the hazard ratio in vaccinated compared to unvaccinated SIREN participants. Vaccine effectiveness will be stratified by vaccine manufacturer. Interaction with demographic factors including sex and ethnicity will be tested. Other analyses will be conducted with the data including estimating the incidence – both asymptomatic and symptomatic – of these pathogens on healthcare workers, and determining their burden, including on time off work. Data will also support the extension into Influenza and RSV of UKHSA models developed to assess nosocomial transmission of SARS-CoV-2.

## Participant Results Reporting

Participants’ access to results depends on the pathway they are enrolled in. Those with local multiplex testing receive their real-time results. Participants in the hybrid pathway receive their SARS-CoV-2 results in real time, but receive a consolidated a report of multiplex results from central laboratories at the end of the study period. Individuals in the postal pathway receive collated results at the study’s end to allow batched testing.

Considering the diverse infection control and occupational health policies at hospital sites, the detection of respiratory viruses, especially when asymptomatic, required careful management. To support sites, a letter from the Chief Investigator was provided. It contained a consensus statement approved by experts in Infectious Diseases, Virology and Infection Control, both within UKHSA and externally. This offered a recommended approach to pathogen detection during the sub-study and facilitated the participation of some sites with vulnerable patient groups.

Participants are informed of the specific results reporting methodology and timeframes before joining the study. These processes were developed through discussions with the participant involvement panel. This information was covered in the PILS.

## Quality assurance and improvement

### Laboratories

All SIREN samples are processed at local, NHS approved laboratories, or for the hybrid and postal pathways, at UKHSA regional laboratories. Laboratories have their own internal quality assurance processes. UKHSA regional laboratories use the Hologic Aptima Panther Fusion assay, a validated assay for detecting of SARS-CoV-2, Influenza A/B and RSV. Quality assurance for the local site laboratories follows the previous SIREN study protocol.

### Distribution of kits

To deliver kits to participants via the postal pathway an external partner was used. A Memorandum of Understanding is in place outlining expected timescales and kit contents. To ensure quality, a vertical audit is performed using a dummy participant with an address and set of barcodes, provided at random in the first batch of participants. The company is unaware that there was a test participant. Receipt of the correctly assembled kit confirms appropriate provision.

### Uploading of data via UKHSA Regional laboratories

As the regional laboratories had not been part of the SIREN study before, dummy data was provided to the UKHSA Regional laboratory to upload to SGSS to ensure that data linkage was successful prior to managing real samples. Real dummy swabs are not provided, tested and uploaded as this is a national surveillance tool.

### Ongoing dialogue

Regular monitoring of three email addresses allows participants, sites, or partners to contact the team directly with any issues. Participants are encouraged to contact the SIREN team if there are any issues with kits. This is monitored daily and any discrepancies or errors are communicated to the distributor.

To ensure any issues are swiftly dealt with there are also multiple meetings held with all partners. Weekly meetings are held with the kit distributors, and fortnightly meetings with the UKHSA regional laboratory network. Weekly drop-ins are available for all sites who have joined winter pressures. Monthly debriefs are held for all SIREN sites, both those who have joined the sub-study and those that have not, providing updates and the chance for feedback. Monthly PI forums welcome all PIs for study updates and feedback. Ad hoc meetings are arranged if any specific issues arise.

### Maximising distribution

Monthly newsletters are created to share key SIREN updates, including new scientific output. Participant webinars are also conducted to present study information to participants address questions or concerns. “Frequently asked Questions” documents are created and shared with participants when sufficient queries are received.

## Patient and public involvement

SIREN has an active, engaged and highly valued participant involvement panel (PIP) who contributed to the design of this sub-study. The PIP meets every 6 weeks and involves a range of participants that were purposively selected to represent the breadth of SIREN study participants. When changes are proposed to the SIREN protocol the central team presents these to the PIP to ensure that participant views are heard. For the Winter Pressures sub study, the PIP committee discussed the proposals and offered suggestions for design and analyses. Discussion themes included how we feed results to participants, the use of a postal pathway, and testing for multiple respiratory pathogens.

## Ethics and dissemination

This protocol was approved by the Berkshire Research Ethics Committee, Health Research Authority (IRAS ID 284460, REC Reference 20/sc0230); all participants gave informed consent prior to enrolment. Participants all gave written consent to join the main study and had inferred consent for additional Winter Pressures testing after receiving the participant information leaflets ahead of their appointments. Participants are advised that they can withdraw at any time and have the option to withdraw their data and samples, or just to withdraw from prospective involvement. Protocol deviations and breaches are monitored by the core SIREN team and any serious breaches reported within a day of occurrence.

The SIREN Study is an urgent priority study and therefore benefits from rapid ethical review. A close working relationship with the ethics committee allowed the approval to be provided for the major protocol amendment in under a month as such REC approval is in place (IRAS ID 284460, REC reference 20/SC/0230) for the SIREN sub-study. The winter pressures sub-study was approved as a major amendment (21). This allowed the expansion of the study test for all respiratory pathogens, including influenza, in addition to SARS-CoV-2. All personal data management is outlined in the SIREN privacy notice and data protection impact assessment.

Dissemination occurs through regular surveillance reports to core policy decision making teams at national expert committees, as well as through peer-reviewed publications and presentations at international meetings and conferences. Participants and NHS sites are invited to webinars and also receive regular newsletters of key study updates, results and information.

**Trial registration number** ISRCTN11041050.

## Discussion

### Strengths

The SIREN winter pressures sub-study showcases the adaptability and leverage of the ambitious research infrastructure created as a pandemic response. With concerns about the burden of respiratory infections including ‘Flu and RSV alongside COVID-19 this winter, we are able to rapidly adapt our study to collect relevant data. This is possible due to the scale of SIREN, the engaged cohort, strong partnerships with NHS sites, surveillance and laboratory capabilities as national public health agencies, and expertise at agile research delivery. Relationships have been primed to address these new questions.^12^ The multi-disciplinary academic partnerships established by SIREN including through the SIREN consortium have meant that we have relationships established which can be deployed and extended. These partnerships, expertise and methodologies can now be extended to working on ‘Flu and RSV, addressing longstanding questions, and aiding workforce planning.

This provides an opportunity to address longstanding unknown questions around ‘Flu, new questions about RSV with vaccinations in the pipeline, and to address workforce planning questions around managing winter pressures. This includes analysis of correlates of protection, an in-depth clinical pathway through the Vibrant study, and statistical modelling looking at nosocomial transmission pathways. Moreover, this includes the ability to look at questions such as the burden of asymptomatic Influenza, the duration of RSV immunity, the impact of a range of respiratory viruses on sick days in the workforce, and real-world vaccine effectiveness this winter.^13-16^ This will also aid the associated symposium of SIREN sub-studies including more detailed characterisation of T-cell response, viral factors associated with escape, and humoral immunity.

The existing SIREN study infrastructure allows for a thorough characterisation of participants. The prospective, longitudinal nature of this study including serum samples before and after confirmed infection enable valuable analysis on correlates of protection, amongst others. The SIREN cohort is engaged and already familiar with the structure of the study, committed to fortnightly PCR testing and surveys. This enables the measurement of asymptomatic and symptomatic infection therefore reducing selection bias. The healthcare worker cohort is a highly exposed cohort, enabling sufficient power for infection and vaccine effectiveness.^17^

The agility of the SIREN study has facilitated a change to the repertoire of respiratory viruses being studied once it became apparent that there would be high rates of influenza and RSV over the winter. Prospective data collection with serial PCR and serum samples regardless of symptoms (pre-event and post event, enables analysis of correlates of protection. This will aid some key developments of previously poorly understood questions, such as the burden of asymptomatic Influenza, the duration of RSV immunity, the impact of a range of respiratory viruses on sick days in the workforce, and real-world vaccine effectiveness this winter. In addition, this will aid the associated symposium of SIREN sub-studies including more detailed characterisation of T-cell response, viral factors associated with escape, and humoral immunity.

These data will help to inform public health policy including potential measures to protect the healthcare workforce and general population.

### Weaknesses

A cohort comprised solely of HCWs may limit the generalisability of the results to the broader UK population. The reduction in the number of NHS sites throughout the study has led to under-representation of certain regions going into this sub-study. The study population is over-represented by certain demographic and professional groups such as female staff, nursing staff and doctors, while under-representing key groups like staff from ethnic minorities, porters and estates. This impairs the generalisability of the data to these groups.

Being a pragmatic sub-study, the unpredictability of the beginning of the Influenza season poses challenges, together with the decentralised delivery model requiring communications and sign up of multiple partners makes it difficult to guarantee an aligned start time.

## Data Availability

All data produced in the present study are available upon reasonable request to the authors

## Acknowledgements

Our special thanks go to the following people who have been enormously helpful: Kevin Ahmed and the rest of the team at the Berkshire Research Ethics Committee. Caroline Williams and colleagues at the NIHR. UKHSA regional lab networks, including Jonathan Turner, Louise Forster, Kat Slater. Panel of UKHSA experts who assisted with the consensus statement. Thanks to the HDR-UK winter pressures funding and to UKHSA for core SIREN study funding. Thanks also to all participants, research team and site staff not mentioned above.

## Collaborators

In addition, we acknowledge the full SIREN study group who were collaborators in study delivery, as listed below: John Northfield, Sean Cutler, Anna Roynon, Maxine Nash, Amanda Dell, Louise Parfitt, Andrea Richards, Andrea Price, Christian Subbe, Caroline Mulvaney Jones, Julia Roberts, Manny Bagary, Nadezda Starkova, Inderpreet Athwal, Louise Hudson, Ashley Jones, Rebecca Chapman, Lucy Booth, Claire Williams, Fiona Adair, April Hawkins, Chinari Subudhi, Scott Latham, Raksha Mistry, Natalie Silva, Abigail Severn, Alejandro Arenas-Pinto, Eva McAlpine, Aran Dhillon, Connor McAlpine, Gosala Gopalakrishnan, Sarah Creer, Eve Etell Kirby, Kim Gray, Joanna Wright, Joely Morgan, Gemma Harrison, Mark Broadhurst, Simon Taylor, Clare McAdam, Natalie Crooks, Stacey Horne, Anna Grice, Nicola Walker, Luke Bedford, Paul Ridley, Alison O’Kelly, Catherine Sinclair, Val Irvine, Elizabeth Boyd, Claire Thomas, Ina Hoad, Tryphena Konala, Judith Radmore, Emily Macnaughton, Sarah Knight, Kim Hulacka, Robert Shorten, Kathryn Hollinshead, Lois Bullen, Robert Shorten, Claire Corless, Sarah Mcloughlin, Bethany Preece, Sarah Baillon, Samantha Hamer, Joanne Edgar, Kelly Moran, Vijayendra Waykar, Charlotte Wesson, Rebecca Rutter, Maureen Williams, Bethany Jones, Russell Coram, Holly Slater, Joanne Jones, Banher Sandhu, Elijah Matovu, Claire Gabriel, Katherine Pagett, Sheron Clarke, Sally Mavin, Sebastien Fagegaltier, Shannon Proctor, Mary Summerscales, Andrew Gibson, Alexandra Cochrane, Dawid Dytmer, Lita Kovina, Grace Davies, Manish Patel, Berni Welsh, Karen Black, Kate Templeton, Sam Donaldson, Andrea Clarke, Jane Crowe, Kadiga Campbell, Barbara Hamilton, Liz Sheridan, Charlotte Barclay, Maxine Ashton, Alison Rodger, Tabitha Mahungu, Debbie Delgado, Julia Vasant, Deborah Howcroft, Sarah Meisner, Abby Rand, Catherine Thompson, Sophia Strong-Sheldrake, Vicky King, Emma Underhill, Kate Seymour, Holly Morgan, Ash Turner, Anne Hayes, Masood Aga, James Pethick, Ashok Dadrah, Thushan de Silva, Helen Shulver, Gareth Stephens, Simon Tazzyman, Mandy Carnahan, Mandy Beekes, Sanal Jose, Jo stickley, Hannah Gibson, Yuri Protaschik, Susan Regan, Alison Campbell, John Day, Swapna Kunhunny, Bernard Hadebe, Paula Harman, Sharon Tysoe, Bridgett Masunda, Nigara Atayeva, Joanne Galliford, Prisca Gondo, Raji Orath Prabakaran, Jane Dare, Qi Zheng, Danielle McCracken, Emmanuel Defever, Ellene Thompson, Lynda Fothergill, Karen Burns, Andrew Higham, Lisa Bishop, Aileen Menzies, Matt Horton, Therese Kelly, Cristina Dragu, David Hilton, Hannah Jory, Penny Harris, Susan Hopkins, Victoria Hall, Jasmin Islam, Ana Atti, Omoyeni Adebiyi, Nick Andrews, Hannah Emmett, Jonathan Broad, Nish Kapirial, Simone Dyer, Sophie Russell, Colin Brown, Joanna Conneely, Paul Conneely, Sarah Foulkes, Nabila Fowles-Gutierrez, Nipunadi Hettiarachchi, Jameel Khawam, Edward Monk, Katie Munro, Andrew Taylor-Kerr, Jean Timeyin, Edgar Wellington, Angela Dunne, Dominic Sparkes, Naomi Platt, Anna Howells, Enemona Adaji, Omolola Akinbami, Palak Joshi, Paola Barbero, Meera Chand, Andre Charlett, Michelle Cole, Claire Neill, Anne-Marie O’Connell, Ferdinando Insalata, Tim Brooks, Maria Zambon, Mary Ramsay, Ayoub Saei, Ezra Linley, Simon Tonge, Ashley Otter, Silvia D’Arcangelo, Cathy Rowe, Amanda Semper, Eileen Gallagher, Robert Kyffin, Kate Howell, Jacqueline Hewson, Iain Milligan, Noshin Sajedi, Davina Calbraith, Caio Tranquillini, Jerry Ye Aung Kyaw, Dianne Corrigan, Lisa Cromey, Lesley Price, Nicole Sergenson, Sally Stewart, Lynne Haahr, Desy Nuryunarsih, Annelysse Jorgenson, Ayodeji Matuluko, Melanie Dembinsky, Desmond Areghan, Alexander Olaoye, Josie Evans, Jennifer Bishop, Jennifer Weir, Laura Dobbie, Andrew Telfer, David Goldberg, David Crossman, Caitlin Plank, Laura Naismith, Ellen De Lacy, Guy Stevens, Susannah Froude, Linda Tyson, Yvette Ellis, Chris Norman.

