## Appendix for "Adapting COVID-19 research infrastructure to capture Influenza and Respiratory Syncytial Virus alongside SARS-CoV-2 in UK healthcare workers Winter 2022/23 and beyond: protocol for a pragmatic sub-study"

### Appendix 1– Regular participant survey

**Welcome**

Thank you for your continued participation in the study, your contribution is critical for informing the UK's response to COVID-19. We will ask you a few short questions about how you have been between 1st to 2nd May.

**Your health over the past 2 weeks:**

Did you have a positive COVID-19 test?

Select all that apply:

- No
- Yes - lateral flow test (LFT)
- Yes - PCR

Please specify the earliest date (between 1st to 2nd May) you tested positive by lateral flow test:

- **Date picker**

Please specify the earliest date (between 1st to 2nd May) you tested positive by PCR:

- **Date picker**

Did you test positive for another respiratory pathogen?

- Yes
- No

(If yes open up tick box)

Select all that apply

- Influenza
  - Influenza A
  - Influenza B
  - Unsure
- Respiratory Syncytial Virus (RSV)
- Non-COVID-19 coronavirus
- Parainfluenza
- Adenovirus
- Human Metapneumovirus
- Rhinovirus/Enterovirus
- *Bordetella pertussis*
- *Chlamydia pneumoniae*
- *Mycoplasma pneumoniae*
- Other respiratory pathogen (please specify)
  - [Free text]

Please specify the earliest date (between 1st to 2nd May) you tested positive:

**Date picker**

Between 1st to 2nd May, have you started to experience any new symptoms in the following list?

Tick all that apply

- A new, continuous cough (coughing a lot for more than an hour, or 3 or more coughing episodes in 24 hours)
- Fever or high temperature (meaning you feel hot to touch on your chest or back)
- Shortness of breath
- Sore throat
- Runny nose
- Headache
- Muscle aches
- Altered sense of smell
- Altered sense of taste
- Extreme fatigue
- Diarrhoea
- Nausea or Vomiting
- Small, itchy red patches, on fingers or toes
- Rash
- Swollen glands

When did these symptoms start?

Approximate dates are fine. Please use the calendar or enter in DD/MM/YYYY, e.g. 21/01/2020.

If you had more than one symptom, enter the date the first symptom started

**Date picker**

Approximately how long did these symptoms last?

If you had more than one symptom, start counting when the first symptom appeared to when the last symptom ended.

- 1 day
- 2 days
- 3 days
- 4 days
- 5 days
- 6 days
- 7 to 14 days (1-2 weeks)
- My symptoms still have not resolved
- I am not sure

In the last 2 weeks have you taken any days off work (sick leave) due to respiratory symptoms?

- Yes
  - Sliding scale 1-14 days
- No

During this period did you seek medical care at hospital for these symptoms?

- Yes, and COVID-19 was confirmed
- Yes, and influenza was confirmed
- Yes, and another respiratory pathogen was confirmed
- Yes, but **no** respiratory pathogen was confirmed
- No

During this period did you receive any treatment for COVID-19?

- Yes
- No

(pops up if yes to having a positive result)

- Nirmatrelvir and ritonavir (Paxlovid)
- Sotrovimab (Xevudy)
- Remdesivir (Veklury)
- Molnupiravir (Lagevrio)
- Dexamethasone
- Casirivimab / imdevimab (Ronapreve)
- Tocilizumab
- Sarilumab
- Baricitinib
- Convalescent plasma
- Palivizumab
- None of the above

**Date picker**

**Your exposures to COVID-19 at work, at home and in the community**

Thinking about the period 1st to 2nd May:

During this period, how often did your job require you to be in close proximity to a suspected or confirmed COVID-19 patients?

- Every shift
- One shift a week or more, but not every shift
- One shift a month or more, but not every week
- One or two shifts in the last 2 weeks
- Never

Between 1st to 2nd May have you had contact with a suspected or confirmed case of COVID-19 at work?

Tick all that apply

- Yes - with a colleague
- Yes - with a patient
- Yes - with bodily fluids
- No

Have you had any contact with suspected or confirmed COVID-19 cases without the PPE specified by your workplace?

Tick all that apply

- Yes – Experienced a breach / contamination whilst wearing PPE
- Yes - Unexpected encounter with a patient or bodily fluids whilst not wearing PPE
- No – I have not had any contact with COVID-19 without PPE specified by your workplace

Between 1st to 2nd May do you believe you were in contact with someone with COVID-19 at home?

Include shared house occupants and visitors if they were living in your household

Tick all that apply

- Yes, suspected case
- Yes, confirmed case (positive PCR or LFT)
- No
- I don't know / I can't remember

Between 1st to 2nd May do you believe you were in contact with someone with COVID-19 in any other setting?

Include visiting friends and family, pubs, restaurants, place of worship and shopping

Tick all that apply

- Yes, suspected case
- Yes, confirmed case (positive PCR or LFT)
- No
- I don't know / I can't remember

When did the person living in your household become unwell with COVID-19 like illness?

If more than one person got ill in the past 2 weeks, give the date the first person became unwell.

Please use the calendar or enter as DD/MM/YYYY, e.g. 21/01/2020.

**Date picker**

During this period, did any household members have to go to hospital due to COVID-19 related symptoms?

- Yes – COVID-19 was suspected/confirmed
- Yes – COVID-19 was not confirmed
- No
- I am not sure

**Vaccines and treatment**

Have you had a COVID-19 vaccination, including a booster?

Include

- Boost dose(s) as the third, fourth or fifth dose

- Vaccines from abroad

Exclude

- Vaccines as part of a trial (there is a question on trials later)

- Yes - I have had a vaccine dose I have not yet reported
- Yes – I already reported my vaccine dose(s) on a previous questionnaire
- No - I have not been vaccinated against COVID-19

Which vaccine dose(s) are you reporting?

- First dose
- Second dose
- Third dose (Booster)
- Fourth dose (Booster)
- Fifth dose (Booster)

Date of fourth (booster) dose:

Date of fifth (booster) dose:

**Date picker**

What is the name/manufacturer of your fourth (booster) dose?

If you do not know the name, please enter 'don't know'

- Pfizer-BioNTech
- Oxford-AstraZeneca
- Moderna (Spikevax)
- Janssen
- Nuvaxoid (Novavax)
- Other

What is the batch number for your fourth (booster) dose?

If unsure, please enter 'don't know'

**Free text**

Have you had a seasonal flu vaccine for 22/23 season?

- Yes - I have had a vaccine I have not yet reported
- Yes - already reported on a previous questionnaire
- No - I have not had a seasonal flu vaccine for 22/23 season
- If yes, what date did you have your flu vaccination?

**Date picker**

What is the name and manufacturer? [open if yes for flu vaccine]

- Quadrivalent Influenza Vaccine, egg-grown (QIVe) (Sanofi Pasteur)
- Influvac sub-unit Tetra, Quadrivalent Influenza Vaccine, egg-grown (QIVe) (Viatris)
- Cell-based Quadrivalent Influenza Vaccine (QIVc) (Seqirus)
- Supemtek, recombinant Quadrivalent Influenza Vaccine (QIVr) (Sanofi Pasteur)
- Adjuvanted Quadrivalent Influenza Vaccine▾ (aQIV) (Seqirus)
- Don’t know

Did you receive the flu vaccine at work? [open if yes for flu vaccine]

- Yes
- No

Travel Information

Between 1st to 2nd May, did you travel abroad?

- Yes
- No
- I don't know / I can't remember

If yes, please only put the name of the country

**Country 1**	**Free text**

**Country 2**	**Free text**

**Country 3**	**Free text**

**Country 4**	**Free text**

**Country 5**	**Free text**

**Postal pathway participants:**

If you are registering a self-swab, please enter number on the barcode below:

If you are not part of the self-swabbing postal pathway, please leave blank.

- Free text box

### Appendix 2: Laboratory flow chart for centralised testing

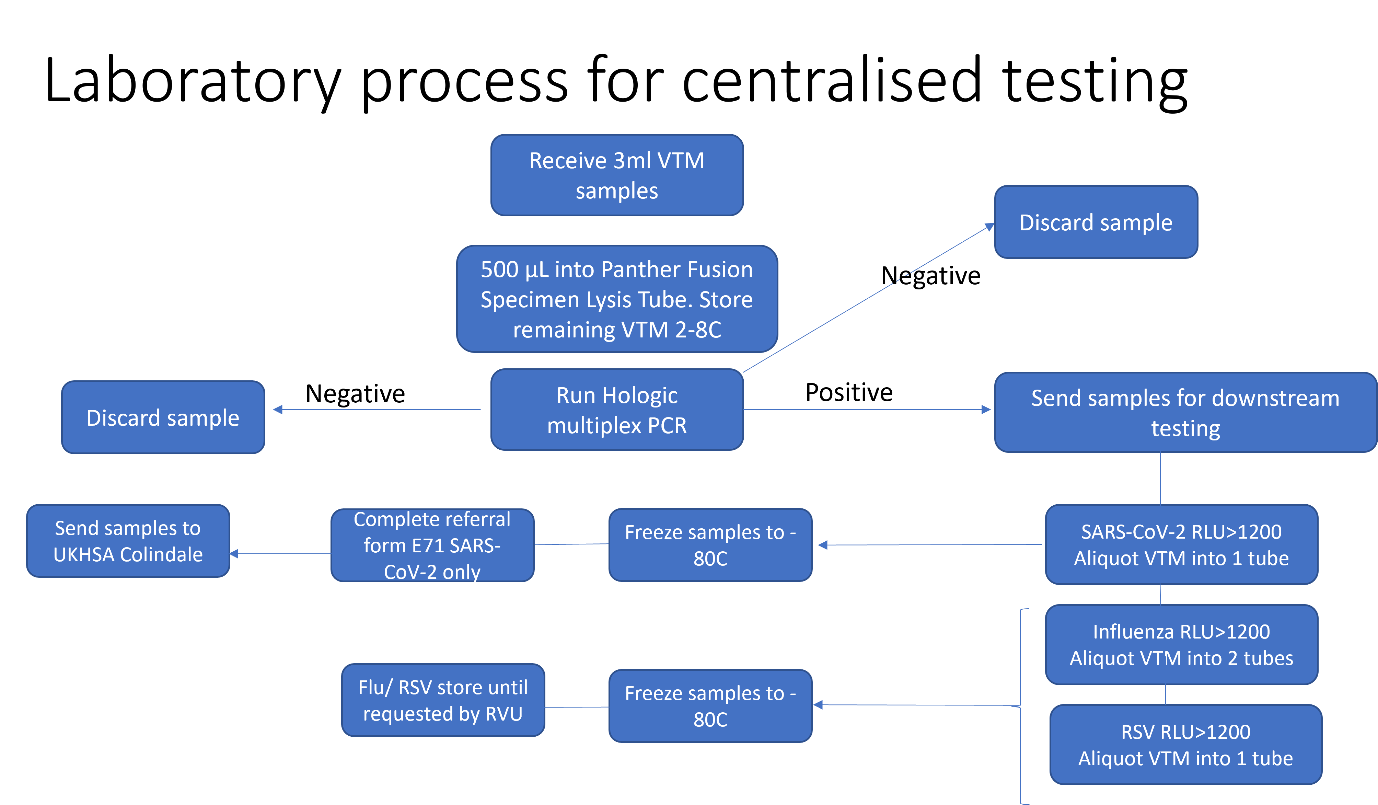

### Appendix 3: referral scheme and protocol between laboratory pathways

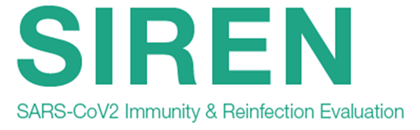

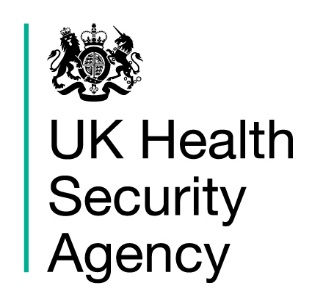

**SOP for referral of samples to the UKHSA regional laboratory network for multiplex testing from SIREN sites offering monoplex testing**

**Pathway for referral of samples from monoplex sites to UKHSA central laboratory network**

The local SIREN team completes the referral form below, including participant name, DOB, SIREN ID, collection date and puts in specimen bag with sample

Results fed back to participant as per current protocol

Sample for SARS-CoV-2 collected and processed in local lab as per current protocol

Participant puts the swab for multiplex testing into a 3ml vial of VTM and into a specimen bag which is collected by the local SIREN team

The local laboratory should pack all of the samples together in a UN3373 compliant box, labelled with UN3373 on the outside for shipping via DX

Specimens should go to the local laboratory as soon as possible but can be kept at 2-8°C for up to 36h

Collect 2 samples:
1 for SARS-CoV-2 (using normal local VTM)
1 for multiplex testing (using 3ml VTM where possible)

The laboratory organises a DX delivery to the XX UKHSA laboratory using DX code XXXXX within 24h of sample receipt

UKHSA Bristol will test the samples for SARS-CoV-2, influenza A + B and RSV and upload results to SGSS. UKHSA SIREN will release these results back to sites at the end of the study

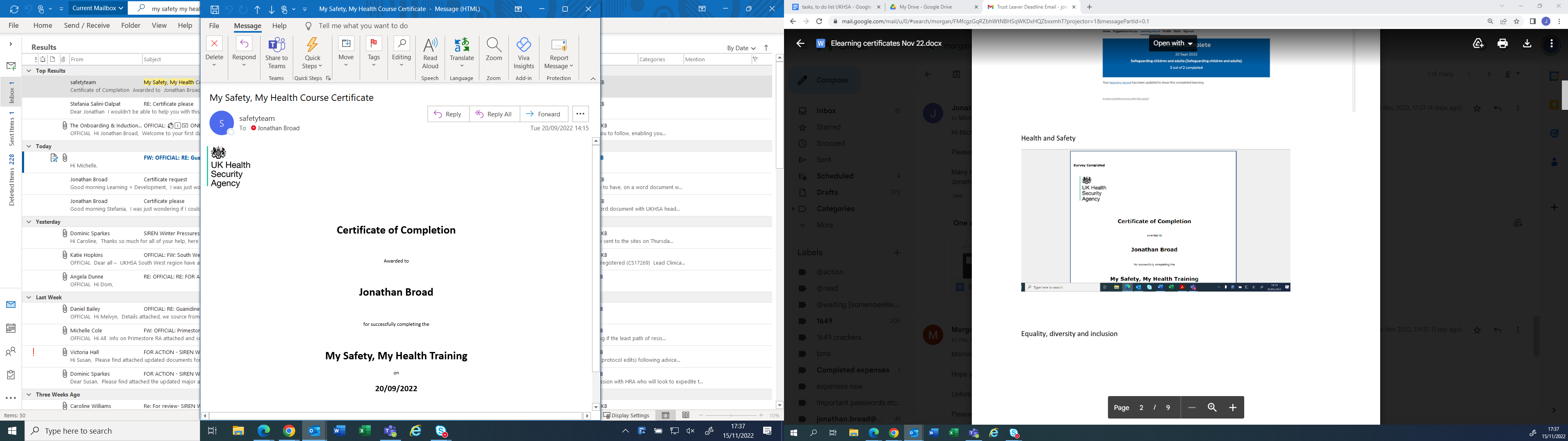

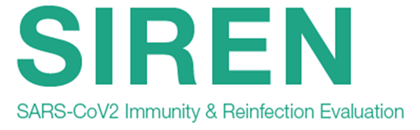

Questions:

**Request form for SIREN: SARS-COV-2, Flu and RSV testing from SIREN sites offering monoplex testing**

**Bristol sites**

| Surname |
| --- |
| Forename |
| Date of birth |
| SIREN ID |
| Swab collection date |
| Please send this sample with the specimen in UN3373 compliant packaging using DX code: XXXXXX  to:  *Address of laboratory*  Please note that results for this sample will be delivered to your site after the end of the study  If there are any queries, please contact us at |

### Appendix 4: Participant information leaflet

**PARTICIPANT INFORMATION LEAFLET: WINTER PRESSURES**

Thank you for your time and commitment to SIREN so far. Every sample and survey you have provided is invaluable and has contributed to our database which continues to answer important questions about SARS-CoV-2 and guide national policy.

**What is the Winter Pressures pathway?**

**The SIREN study has helped to answer many questions about SARS-CoV-2 infection and COVID-19 vaccination. Many of these questions remain unanswered for other respiratory infections that have been around for much longer, in particular influenza (also known as ‘flu’).**

**To help answer some of these questions, the SIREN Winter Pressures pathway will run in certain SIREN sites. The fortnightly swabs (PCR samples) that are already being collected will be tested on a multiplex PCR assay, which simultaneously tests for more respiratory pathogens, instead of just testing for COVID-19. The panel of respiratory infections varies according to which system each laboratory has but will include:**

- **SARS-CoV-2**
- **Influenza A**
- **Influenza B**
- **+/- other respiratory infections, such as Respiratory Syncytial Virus (RSV) or the common cold**

**We will also collect information about the brand of influenza vaccine that each participant has received to compare the effectiveness of the different vaccines available.**

**Why is this important?**

**This year, Australia has reported the highest levels of influenza in the last 5 years following the relaxation of social distancing measures. This is the first winter with minimal social distancing measures, therefore we expect that we may experience a similar pattern in the United Kingdom. This is the first winter that we anticipate both influenza and COVID-19 infections causing sickness in healthcare workers.**

**The SIREN study, with its high number of engaged participants undergoing regular testing, has proven that it is well placed to answer questions on SARS-CoV-2 infection and vaccine effectiveness. We can answer similar questions for influenza without having to collect extra samples from participants. It is unlikely that there will be a cohort of such participants, or sites to help with data collection, in future years; this is the ideal time to explore important influenza research questions.**

**Will my follow up and sampling be the same?**

**Yes, many participants will have the same frequency of PCRs, serology and surveys as currently. Those sites who are offering quarterly serology will be asked to increase to monthly where possible, to help answer questions about the effect of vaccination on the immune response.**

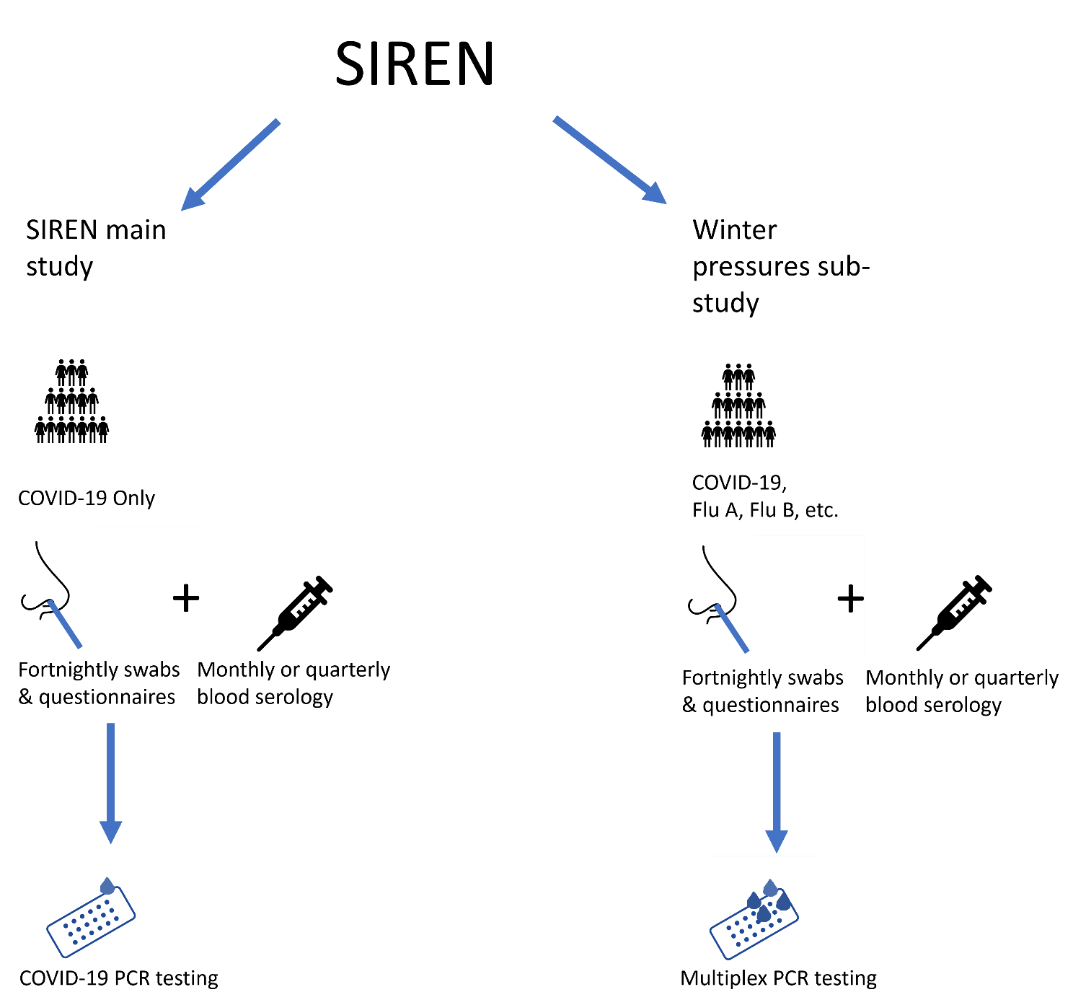
**The below figure depicts the two pathways that will run in parallel over this winter. The main SIREN study demonstrates the structure of the current testing, which will be continued by some sites, whilst the winter pressures pathway sites will process the fortnightly PCR samples using a multiplex testing platform. You will be informed which pathway your site has decided to offer before any change to your testing.**

**All data management and information governance procedures remain the same.**

**What do you want me to do now?**

If you are happy to continue you do not need to do anything except attend your appointments as scheduled. There is no paperwork to complete. If your site decides to use the multiplex assay you will be informed of this by your SIREN site team, including the pathogens being tested. Attending your appointments as usual will count as consent that you are happy to provide PCR swabs and blood samples for this sub study.

As it is logistically challenging for a site to offer more than one testing pathway sites will offer either multiplex PCR or COVID-19 PCR alone (monoplex). If you do not wish to have the additional respiratory PCR testing but are at a site performing multiplex for other respiratory infections, please discuss this with your site team for consideration of a transfer or withdrawal.

**We will ask all participants in SIREN sites to record the brand name and type of influenza vaccination received in their SIREN follow-up survey**. You might find it helpful to photograph the box or retain the leaflet from the ‘flu clinic so that you can provide this information when asked. This is particularly important as the National Immunisation Database may not always record the exact vaccination that each person has received, and some brands produce more than one type of ‘flu vaccine.

Finally, please keep a record of the number of days that you have taken off work sick for respiratory illnesses, to ensure this is completed in the survey.

**What happens if I test positive for SARS-CoV-2, Influenza or one of the other pathogens?**

Different healthcare trusts vary according to their Occupational Health and Infection Control policies. Individuals with respiratory symptoms should take precautions and follow local policy regarding time off work, regardless of these results. In those who are asymptomatic the risk of transmitting to others is low. However, certain precautions may be sensible based on the patients you have contact with. For example, this might include mask wearing or avoiding high risk patients. We advise vigilance for symptoms following a positive test. If sites only perform SARS-CoV-2 testing locally and send away the samples for the other pathogens, there will be a delay in the multiplex results reaching participants. You will be informed if this delay will apply to your site.

A letter from the UKHSA has been sent to hospital trusts regarding participants involved in SIREN to suggest these and other measures. Certainly, participants should not be penalised for diagnoses or time off work as a result as being involved in this study, if support is required, please do contact us.

**What are the benefits to me?**

The study will not benefit you directly, but your participation will help provide important information about SARS-CoV2, influenza and other respiratory infections among staff working in healthcare organisations and provide a stronger evidence base to inform national guidance and policy. At the end of the study, the overall results will be published in national reports and longstanding questions about influenza answered with these data. Participants will receive results from their sites for all these pathogens via the usual route.

**What happens if my site does not join the winter pressures sub-study?**

If your site does not choose to join the winter pressures sub-study, you will continue the SIREN study as it currently stands. You will continue fortnightly surveys, fortnightly PCR swab tests, and monthly serology tests (or quarterly if your site is unable to offer this). Your PCR swab samples will continue to be used for COVID-19 testing, which continue to contribute to the wealth of information coming from the SIREN study. We may ask your site to send your samples to the central lab at a later stage to be tested for other respiratory infections and we would return these results to your local SIREN site. We will inform you that we have requested this from your site at that time.

**What if I change my mind?**

If you no longer want to be involved, you can withdraw from the study at any time by visiting the SIREN website and following the link to withdraw from the study: <https://snapsurvey.phe.org.uk/siren/>. Withdrawal is not complete until you have completed the survey which will be sent you to via email or text message, that you can access at this link.

**Who has reviewed the study?**

All research in the NHS is looked at by an independent group of people, called a Research Ethics Committee, to protect participants’ interests. This study has been reviewed and given favourable opinion by Berkshire Research Ethics Committee.

Within SIREN we have a Participant Involvement Panel (PIP), composed of a selection of SIREN participants who meet regularly to review and give feedback on study developments. Members are recruited from a range of professional backgrounds, geographic regions and demographic groups to represent the diversity of the SIREN cohort. The winter pressures pathway has been presented and discussed with the PIP, with positive feedback and suggestions, which have guided the protocol amendment and study documents.

**Who should I contact with feedback or questions?**

***Thank you for reading this information and continuing to play your part in SIREN.***

### Appendix 5 Participant information leaflet – postal pathway

**PARTICIPANT INFORMATION LEAFLET: WINTER PRESSURES SUBSTUDY - POSTAL SWABS PATHWAY**

Thank you for your time and commitment to SIREN so far.  Every sample and survey you have provided is invaluable and has contributed to our database which continues to answer important questions about SARS-CoV-2 and guide national policy.

**What is the Winter Pressures pathway?**

The SIREN study has helped to answer many questions about SARS-CoV-2 infection and COVID-19 vaccination.  Many of these questions remain unanswered for other respiratory infections that have been around for much longer, in particular influenza (also known as ‘flu’).

To help answer some of these questions, the SIREN Winter Pressures sub study will run over this winter. There will be two pathways in this sub study:

- Some sites will switch to running their PCR samples on a multiplex panel
- Participants from sites that have closed out or are unable to offer multiplex testing can join a postal swab pathway.

The fortnightly swabs (PCR samples) will be tested on a multiplex PCR assay, which simultaneously tests for more respiratory pathogens, instead of just testing for COVID-19.  The panel of respiratory infections varies according to which system each laboratory has but will include:

- SARS-CoV-2
- Influenza A
- Influenza B
- +/- other respiratory infections, such as Respiratory Syncytial Virus (RSV) or the common cold

We will also collect information about the brand of influenza vaccine that each participant has received to compare the effectiveness of the different vaccines available.

**Why is this important?**

This year, Australia has reported the highest levels of influenza in the last 5 years following the relaxation of social distancing measures. This is the first winter with minimal social distancing measures, therefore we expect that we may experience a similar pattern in the United Kingdom.  This is the first winter that we anticipate both influenza and COVID-19 infections causing sickness in healthcare workers.

The SIREN study, with its high number of engaged participants undergoing regular testing, has proven that it is well placed to answer questions on SARS-CoV-2 infection and vaccine effectiveness.  We can answer similar questions for influenza without having to collect extra samples from participants.  It is unlikely that there will be a cohort of such participants, or sites to help with data collection, in future years; this is the ideal time to explore important influenza research questions.

**Will my follow up and sampling be the same?**

It will be very similar. Participants will have the same frequency of PCRs and surveys as currently, however, in the postal pathway participants **will not** undergo serology testing, as this relies on access to a local site with capacity.  Nevertheless, there are many research questions that can still be answered without these serology samples.

The below figure depicts the structure of the SIREN study over this winter. The main SIREN study demonstrates the structure of the current testing, which will be continued by some sites, whilst the winter pressures pathways will utilise multiplex testing at sites or via a postal swab system.  You have received this leaflet as you are eligible to join winter pressures postal swab pathway, highlighted by the red box.

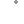

***Please note – some of the swab tubes may contain a chemical called Guanidine, which is an irritant to skin and eyes and caution should be taken to follow the hazard advice as per the user instructions.*** Some of the kit components will reach their expiry date during the study period, although the manufacturers provided an exemption letter that permits usage 6 months beyond expiry date.

**Is there any change to the data management and information governance procedures?**

All data management and information governance procedures remain largely the same, however the names and addresses of participants in this pathway will be shared with an approved company that is contracted by UKHSA to help send kits, who will use this minimal information to distribute the postal packs.  No other SIREN data will be shared with this partner. This is included in our privacy statement: <https://snapsurvey.phe.org.uk/siren/winterpressures/siren_privacy_notice.pdf>. This does not affect any previous consent statements.

**What do you want me to do now?**

If you would like to join this SIREN winter pressures sub study and are able to provide fortnightly PCRs via post, along with fortnightly questionnaires, we ask you to complete the electronic consent form.  Please note that when we have enough participants, we will close the survey. You will shortly after receive the swab packs via post, along with instructions and labels for the samples, and prepaid packaging.  The success of this study will rely on regular samples being sent, so please only join if you are willing to self-swab every two weeks and are able to post the samples promptly after swabbing. Please ensure you are reporting the swabs at the same time as your survey, and please remember to stick barcodes onto the sample, the form and to register them on your survey at the same time.

**We will ask all participants in SIREN sites to record the brand name and type of influenza vaccination received in their SIREN follow-up survey**.  You might find it helpful to photograph the box or retain the leaflet from the ‘flu clinic so that you can provide this information when asked.  This is particularly important as the National Immunisation Database may not always record the exact vaccination that each person has received, and some brands produce more than one type of ‘flu vaccine.

Finally, please keep a record of the number of days that you have taken off work sick for respiratory illnesses, to ensure this is completed in the survey.

**What happens if I test positive for SARS-CoV-2, Influenza or one of the other pathogens?**

Due to the centralised testing that will be used for this postal pathway, **you** **will receive the results of all tests at the end of the study** as the centralised laboratory and SIREN team do not have capacity to perform and return results in real-time.  This is purely a research study, therefore we ask you to follow local guidelines regarding testing and isolation in the event of symptoms, as if you were not part of the study.

**What are the benefits to me?**

The study will not benefit you directly, but your participation will help provide important information about SARS-CoV2, influenza and other respiratory infections among staff working in healthcare organisations and provide a stronger evidence base to inform national guidance and policy. At the end of the study, the overall results will be published in national reports and longstanding questions about influenza answered with these data.

**What if I change my mind?**

If you decide to join this study but subsequently no longer want to be involved, you can withdraw from the study at any time. If you wish to stop this additional postal testing please email us at email us at. You can withdraw completely from the entire study by visiting the SIREN website and following the link to withdraw from the study: <https://snapsurvey.phe.org.uk/siren/>.  Withdrawal is not complete until you have completed the survey which will be sent you to via email or text message, that you can access at this link.  Please note that withdrawal is not necessary if you do not wish to join this study; you can either ignore this communication or select ‘I do not wish to join the Winter pressures postal pathway’ on the participant survey.

**Who has reviewed the study?**

All research in the NHS is looked at by an independent group of people, called a Research Ethics Committee, to protect participants’ interests. This study has been reviewed and given favourable opinion by Berkshire Research Ethics Committee.

Within SIREN we have a Participant Involvement Panel (PIP), composed of a selection of SIREN participants who meet regularly to review and give feedback on study developments. Members are recruited from a range of professional backgrounds, geographic regions and demographic groups to represent the diversity of the SIREN cohort. The winter pressures pathway has been presented and discussed with the PIP, with positive feedback and suggestions, which have guided the protocol amendment and study documents.

**Who should I contact with feedback or questions?**

***Thank you for reading this information and continuing to play your part in SIREN.***

### Appendix 6: Participant information leaflet for centralised pathway

**PARTICIPANT INFORMATION LEAFLET: WINTER PRESSURES SUBSTUDY – SITES USING CENTRAL LABORATORIES**

Thank you for your time and commitment to SIREN so far.  Every sample and survey you have provided is invaluable and has contributed to our database which continues to answer important questions about SARS-CoV-2 and guide national policy.

What is the Winter Pressures pathway?

The SIREN study has helped to answer many questions about SARS-CoV-2 infection and COVID-19 vaccination.  Many of these questions remain unanswered for other respiratory infections that have been around for much longer, in particular influenza (also known as ‘flu’).  To help answer some of these questions, the SIREN Winter Pressures sub study will run over this winter. There will be two site-based pathways in this sub-study:

- Some sites will switch to running their PCR samples for COVID-19, influenza A+B and RSV locally
- Some sites will switch to running their COVID testing locally, but their other viruses in a centralised UKHSA laboratory.

The fortnightly swabs (PCR samples) will be tested on a multiplex PCR assay, which simultaneously tests for more respiratory pathogens, instead of just testing for COVID-19.  The panel of respiratory infections varies according to which system each laboratory has but will include:

- SARS-CoV-2
- Influenza A
- Influenza B
- +/- other respiratory infections, such as Respiratory Syncytial Virus (RSV) or the common cold

If you have been vaccinated, we will also collect information about the brand of influenza vaccine that each participant has received to compare the effectiveness of the different vaccines available.

**Why is this important?**

This year, Australia has reported the highest levels of influenza in the last 5 years following the relaxation of social distancing measures. This is the first winter with minimal social distancing measures, therefore we expect that we may experience a similar pattern in the United Kingdom.  This is the first winter that we anticipate both influenza and COVID-19 infections causing sickness in healthcare workers.

The SIREN study, with its high number of engaged participants undergoing regular testing, has proven that it is well placed to answer questions on SARS-CoV-2 infection and vaccine effectiveness.  We can answer similar questions for influenza without having to collect extra samples from participants.  It is unlikely that there will be a cohort of such participants, or sites to help with data collection, in future years; this is the ideal time to explore important influenza research questions.

**Will my follow up and sampling be the same?**

Yes, many participants will have the same frequency of PCRs, serology and surveys as currently.  Those sites who are offering quarterly serology will be asked to increase to monthly where possible, to help answer questions about the effect of vaccination on the immune response.  Your site may ask you to provide an additional swab, one for local COVID-19 testing and the other for the additional testing of Influenza A, Influenza B and RSV. If you do not want to perform an additional test liaise with your site and you can remain providing a single swab for COVID-19.

**Is there any change to the data management and information governance procedures?**

All data management and information governance procedures remain largely the same, however your additional test results will be performed at an approved centralised laboratory. This is included in our privacy statement: [**https://snapsurvey.phe.org.uk/siren/winterpressures/siren_privacy_notice.pdf**](https://snapsurvey.phe.org.uk/siren/winterpressures/siren_privacy_notice.pdf). Your consent to this does not affect any previous consent statements.

**What do you want me to do now?**

If you are happy to continue you do not need to do anything except attend your appointments as scheduled.  There is no paperwork to complete.  If your site decides to use the multiplex centralised test you will be informed of this by your SIREN site team, including the pathogens being tested. Attending your appointments as usual will count as consent that you are happy to provide PCR swabs and blood samples for this sub study.

Your site may offer this testing via performing an additional swab or via using leftovers from the sample.  As it is logistically challenging for a site to offer more than one testing pathway sites will largely offer either multiplex PCR or COVID-19 PCR alone (monoplex). If you do not wish to have the additional respiratory PCR testing but are at a site performing multiplex for other respiratory infections, please discuss this with your site team for consideration of withdrawal or whether they can offer you the COVID-19 PCR only pathway.

We will ask all participants in SIREN sites to record the brand name and type of influenza vaccination received in their SIREN follow-up survey. If you do not know this or if you are unvaccinated, you can still take part. You might find it helpful to photograph the box or retain the leaflet from the ‘flu clinic so that you can provide this information. This is important as the National Immunisation Database may not always record the exact vaccination that each person has received, and some brands produce more than one ‘flu vaccine.

Finally, please keep a record of the number of days that you have taken off work sick for respiratory illnesses, to ensure this is completed in the survey.

**What happens if I test positive for SARS-CoV-2, Influenza or one of the other pathogens?**

Different healthcare trusts vary according to their Occupational Health and Infection Control policies. Individuals with respiratory symptoms should take precautions and follow local policy regarding time off work, regardless of any results. Local policies should be followed if you test positive for COVID-19. Due to the centralised testing that will be used for the other viruses in this pathway, **you** **will receive the results of all other tests at the end of the study** as the centralised laboratory and SIREN team do not have capacity to perform and return results in real-time.  This is purely a research study, therefore we ask you to follow local guidelines regarding testing and isolation in the event of symptoms, as if you were not part of the study.

**What are the benefits to me?**

The study will not benefit you directly, but your participation will help provide important information about SARS-CoV2, influenza and other respiratory infections among staff working in healthcare organisations and provide a stronger evidence base to inform national guidance and policy. At the end of the study, the overall results will be published in national reports and longstanding questions about influenza answered with these data.

**What if I change my mind?**

If you decide to join this study but subsequently no longer want to be involved, you can withdraw from the study at any time. If you wish to stop the additional testing please speak to your local SIREN team about whether they can offer you the COVID-19 only pathway or by emailing us at:. You can withdraw completely from the entire study by visiting the SIREN website and following the link to withdraw from the study: <https://snapsurvey.phe.org.uk/siren/>.  Withdrawal is not complete until you have completed the survey which will be sent you to via email or text message, that you can access at this link.

**Who has reviewed the study?**

All research in the NHS is looked at by an independent group of people, called a Research Ethics Committee, to protect participants’ interests. This study has been reviewed and given favourable opinion by Berkshire Research Ethics Committee. Within SIREN we have a Participant Involvement Panel (PIP), composed of a selection of SIREN participants who meet regularly to review and give feedback on study developments. Members are recruited from a range of professional backgrounds, geographic regions and demographic groups to represent the diversity of the SIREN cohort. The winter pressures pathway has been presented and discussed with the PIP, with positive feedback and suggestions, which have guided the protocol amendment and study documents.

**Who should I contact with feedback or questions?**

***Thank you for reading this information and continuing to play your part in SIREN.***

### Appendix 7: Swab instructions

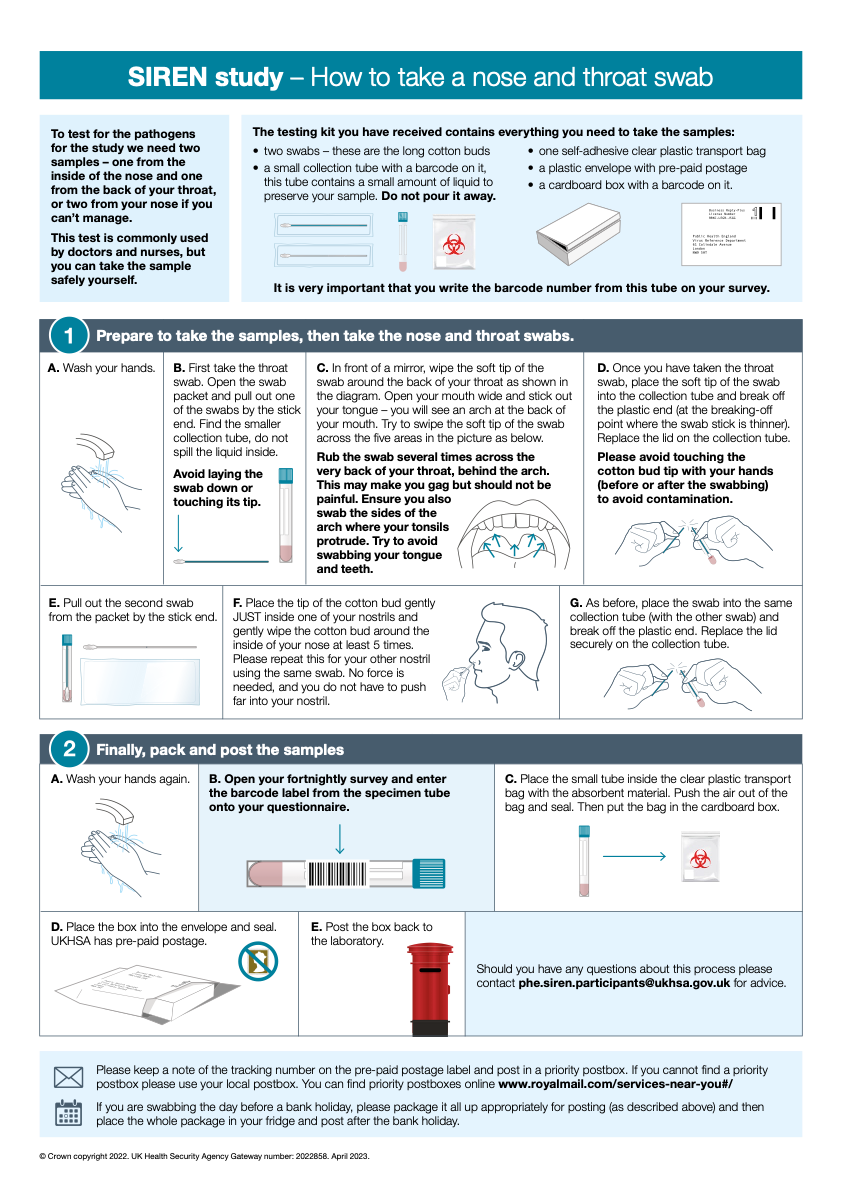
